# A Cardiac-specific CT Foundation Model for Heart Transplantation

**DOI:** 10.1101/2025.08.14.25333618

**Authors:** Hanwen Xu, Addie Woicik, Sanaz Asadian, Junbo Shen, Zhengyan Zhang, Ali Nabipoor, J. Peter Musi, Jeffrey Keenan, Maziar Khorsandi, Bassel Al-Alao, Ioannis Dimarakis, Hamid Chalian, Yiing Lin, Daniel Fishbein, Jay Pal, Sheng Wang, Shin Lin

## Abstract

Heart failure is a major cause of morbitidy and mortality, with the severest forms requiring heart transplantation. Heart size matching between the donor and recipient is a critical step in ensuring a successful transplantation. Currently, a set of equations based on population measures of height, weight, sex and age, viz. predicted heart mass (PHM), are used but can be improved upon by personalized information from recipient and donor chest CT images.

Here, we developed *GigaHeart*, the first heart-specific foundation model pretrained on 180,897 chest CT volumes from 56,607 patients. The key idea of *GigaHeart* is to direct the foundation model’s attention towards the heart by contrasting the heart region and the entire chest, thereby encouraging the model to capture fine-grained cardiac features.

*GigaHeart* achieves the best performance on 8 cardiac-specific classification tasks and further, exhibits superior performance on cross-modal tasks by jointly modeling CT images and reports. We similarly developed a thorax-specific foundation model and observed promising performance on 9 thorax-specific tasks, indicating the potential to extend *GigaHeart* to other organ-specific foundation models.

More importantly, *GigaHeart* addresses the heart sizing problem. It avoids oversizing by correctly segmenting the sizes of hearts of donors and recipients. In regressions against actual heart masses, our AI-segmented total cardiac volumes (TCVs) has a 33.3% *R*^2^ improvement when compared to PHM. Meanwhile, *GigaHeart* also solves the undersizing problem by adding a regression layer to the model. Specifically, *GigaHeart* reduces the mean squared error by 57% against PHM. In total, we show that *GigaHeart* increases the acceptable range of donor heart sizes and matches more accurately than the widely used PHM equations.

In all, *GigaHeart* is a state-of-the-art, cardiac-specific foundation model with the key innovation of directing the model’s attention to the heart. *GigaHeart* can be finetuned for accomplishing a number of tasks accurately, of which AI-assisted heart sizing is a novel example.

## Main

Of the more than 6.7 million individuals with heart failure in the U.S. [1], approximately 4500 with the severest forms undergo heart transplantation each year [2]. This procedure is exceedingly resource intensive, and a shortage of donor organs persists, limiting the widespread applicability of this therapy, despite excellent clinical outcomes. Thus, appropriate donor-recipient matching is critical for distributing this limited resource to the most suitable patients. Matching the size of the donor heart to the intended recipient is an integral part of the donor evaluation process [3].

Inaccurate heart size estimation can lead to poor post-transplant outcomes. Undersizing the donor heart can lead to graft failure [4] while oversizing may lead to decreased survival stemming from complications related to cardiac compression or delayed chest closure [5]. In the past, clinicians undertook size matching between the donor and recipient by comparing weigh, height, or a combination of the two, which later proved to be clinically irrelevant [5]. More recently, clinicians use a set of equations to calculate the predicted heart mass (PHM) for both the donor and recipient based on age, height, weight and sex [6]. These calculations have been shown to accurately predict cardiac mass, but the potential for significant residual errors exists which may result in adverse clinical outcomes.

Chest CT scans are almost always available for both the donor and recipient; these studies could make size matching more precise and individualized than equations dervied from population level data, such as PHM. Cardiac MRIs or dedicated ECG-gated cardiac CTs yield the most accurate measures of heart volume but cannot be practically obtained in the transplant setting. This is because these studies require extra technical steps and the administration of intravenous contrast, which may harm the donor kidney. Therefore, it is critical to develop methods that work with the chest CTs, which are commonly obtained in the clinical setting of heart transplant.

Current approaches for estimating heart size from CT scans rely heavily on manual or semi-automated segmentation, which is logistically impractical for the heart transplantation workflow. For example, having a radiologist or imaging technician on standby to perform segmentation for donor heart offers, which may appear at any time of the day or night, is not feasible for most centers. Developing machine learning models for automating heart size estimation from CT images is also challenging due to the small amount of annotated data. Recently, foundation models, which leverage large-scale unlabeled data to generate high-quality, transferable features, have emerged as a promising paradigm for this type of scenario [7, 8, 9, 10, 11, 12, 13, 14, 15]. CT-based foundation models such as MedVersa [16], CT-CLIP [17], and Merlin [18] have shown strong performance across a broad range of tasks. However, these models are typically trained on general chest or abdominal scans without specific adaptation to cardiac anatomy or cardiac-specific applications. As a result, their utility and generalizability to heart-specific tasks remain uncertain, especially for cardaic sizing, which requires precise modeling of subtle anatomical features surrounding the heart.

Moreover, a unique challenge in heart sizing is to estimate the normal-sized heart of the recipient as if they never had disease. In particular, as part of the disease process, the recipient heart is likely to enlarge. To date, we are not aware of any program which uses patient CT scans to impute these volumes.

Here, we present *GigaHeart*, a novel organ-specific foundation model designed to improve performance on cardiac-specific tasks, especially heart sizing, using general chest CT data. Our key technical idea is to direct the foundation model’s attention toward the heart by contrasting representations between the heart region and the entire chest, thereby encouraging the model to capture fine-grained cardiac features and generalize more effectively to downstream heart-related applications. Specifically, our approach begins by training a base CT foundation model on 180,897 chest CT volumes, comprising 60,320,854 individual, 2D scans from 56,067 patients. We then train a heart segmentation model using 2,262 2D expert-annotated heart masks from 70 CT volumes and use it to extract 8,656,368 heart-region views from the original CT volumes. We then continue training the base foundation model by forcing it to focus only on these heart-region views, effectively adapting it for heart-focused tasks.

*GigaHeart* represents the first attempt to develop a fine-grained, cardiac-specific foundation model using general chest CT data. We demonstrated *GigaHeart*’s versatility across a wide range of cardiovascular tasks. First, *GigaHeart* outperforms existing CT foundation models on 8 cardiac-specific classification tasks and shows superior generalizability in both cross-device and cross-cohort evaluations. Moreover, *GigaHeart* also proves effective in multi-modal settings, achieving state-of-the-art performance in zero-shot classification, cross-modal retrieval, and visual question answering. More importantly, *GigaHeart* attains superior performance on heart sizing as validated in multiple ways over the current standard-of-care estimation with PHM. Finally, we showed the broader applicability of *GigaHeart*by similarly developing a thorax-specific foundation model using chest CT data. This model also achieves significant improvement over others on 9 thorax-specific classification tasks, multi-modal applications and CT-based segmentation, indicating the versatility of our framework to develop organ-specific foundation models. To facilitate future research, we will open-source our model, code and release the pre-trained CT images used to develop *GigaHeart*.

## Results

### Overview of *GigaHeart*

To develop a foundation model that is capable of cardiac relevant abnormality detection and heart size matching from chest CT images, we propose a novel heart-aware pretraining method that combines both the standard state-of-the-art self-supervised learning approach DINOv2 [19] as well as supervision signals from experts to direct the model’s attention to cardiovascular structures of special interest within the image volumes (**Fig. 1A**). To this end, we curated a large scale dataset comprising 180,897 chest CT volumes obtained from UW Medicine constituting, to our knowledge, the largest chest CT dataset assembled to date.

**Figure 1:**
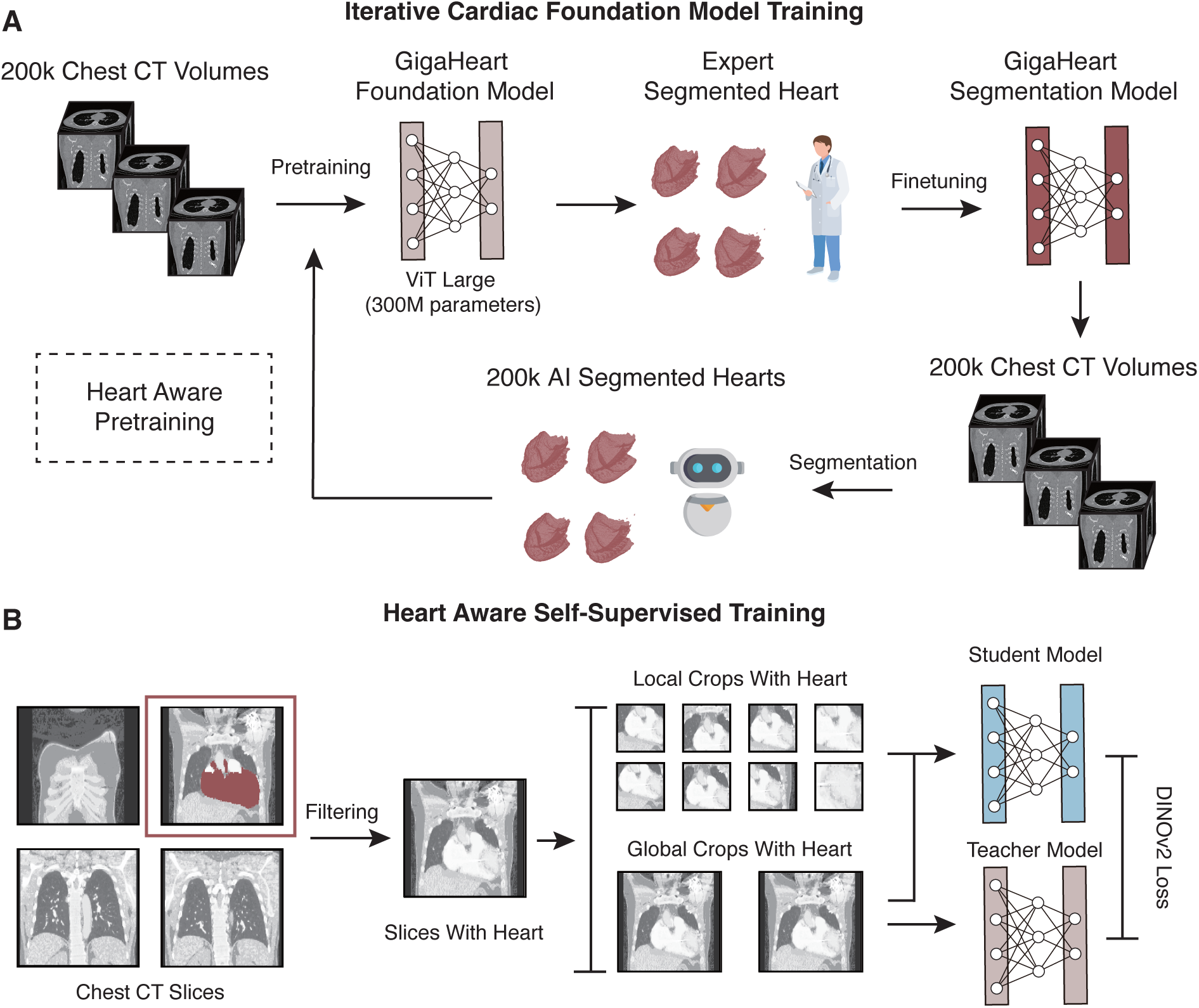
Overview of *GigaHeart*. **A,** *GigaHeart* is a chest CT foundation model tailored to cardiovascular relevant tasks through an iterative expert-in-the-loop training pipeline. The *GigaHeart* model is a large vision transformer (ViT) composed of 300 million parameters. Initially, it undergoes standard DINOv2 pretraining using 180,897 unlabeled chest CT volumes. Subsequently, the pretrained model is finetuned using expert-segmented heart masks, resulting in the *GigaHeart* segmentation model. This segmentation model then autonomously segments all the chest CT volumes, generating high-quality heart masks. *GigaHeart* is further refined through a heart-aware, DINOv2, second-stage pretraining step. **B,** The random cropping in DINOv2 is replaced with heart-aware cropping, in which each generated crop must contain the whole heart or part of the heart. This adaptation focuses the model’s attention on this organ, thus improving the model’s capability of extracting cardiovascular related CT patterns.

The core insight of our work is that standard self-supervised learning methods, derived primarily from general-domain datasets such as ImageNet [20], are suboptimal for addressing specialized medical tasks like heart size matching. Specifically, standard DINOv2 pretraining equally weights all image patches and regions through its multi-cropping DINO loss and masked-patch reconstruction iBOT loss, inadvertently diluting crucial signals from cardiovascular regions. To address this limitation, we first initialize *GigaHeart* with conventional DINOv2 pretraining. Following initial convergence, we leverage a small dataset of 70 clinician-annotated heart segmentation masks to fine-tune the pretrained model into an accurate AI segmentation tool. This tool subsequently generates heart segmentation masks across our entire pretraining dataset, enabling a targeted, heart-aware adaptation of the DINOv2 training process. Crucially, we integrate these generated heart masks into our pretraining pipeline by constraining every random crop to either fully encompass the heart or include portions of it (**Fig. 1B**). This straightforward yet powerful adaptation significantly enhances the model’s ability to focus specifically on cardiovascular features, demonstrating notable effectiveness in improving cardiac region awareness.

### *GigaHeart* accurately predicts cardiac-specific abnormalities

We first evaluated the performance of *GigaHeart* on predicting cardiac-specific abnormalities by separately finetuning it to detect 7 cardiovascular abnormalities on two independent benchmarks (**Fig. 2**). We compared its performance against the latest CT foundation models including Merlin [18] and SegVol [21] as well as foundation models in the general domain, such as DINOv2 [19] and 3D ResNet-50 [22, 23]. *GigaHeart* substantially outperforms the competing approaches and achieves the state-of-the-art performance in detecting 5 out of 6 abnormalities on RadChestCT (**Fig. 2A**, Supplementary Fig. 3), indicating its superior performance on cardiac-specific tasks by focusing on heart-region views. On another benchmark CT-RATE, *GigaHeart* further achieves the best performance on 2 out of 2 abnormalities (**Fig. 2A**, Supplementary Fig. 3). To understand the superiority of *GigaHeart*, we performed ablation studies removing the heart-aware training component. We observe significant performance drop, highlighting the effectiveness of continual training on heart-region views (Supplementary Fig. 1).

**Figure 2:**
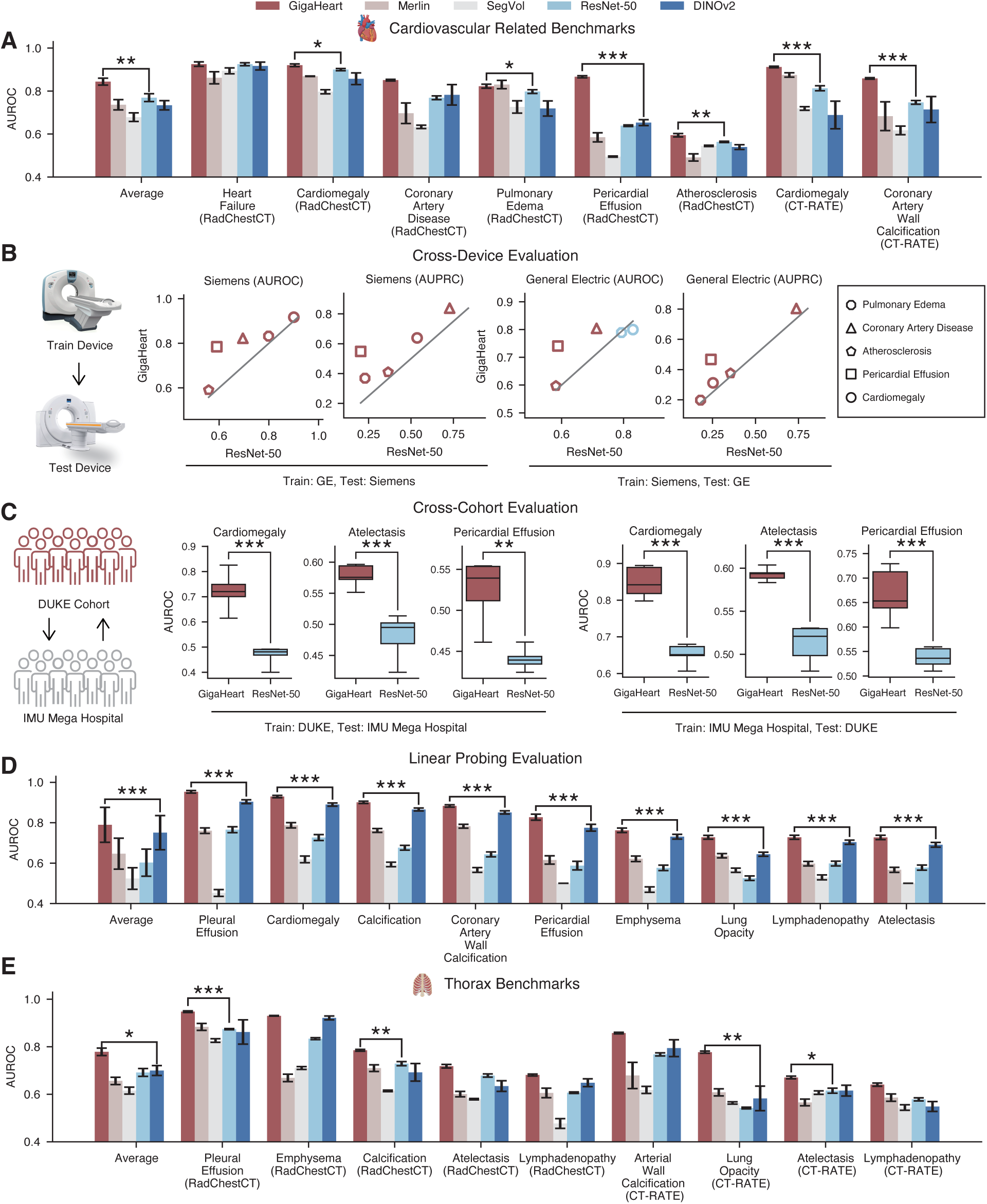
Evaluation of cardiovascular and thorax relevant benchmarks. **A, E** Bar plots comparing *GigaHeart* and competing methods on cardiovascular and thorax-related benchmarks in terms of area under the receiver-operator curve (AUROC). **B,** Scatter plots comparing the performance between *GigaHeart* and ResNet-50 in terms of AUROC and the area under the precision-recall curve (AUPRC) on cross-device evaluations. Each dot indicates one cardiovascular-related task. **C,**Boxplots comparing the performance between *GigaHeart* and ResNet-50 in terms of AUROC and AUPRC on cross-cohort evaluations. **D,** Barplots comparing AUROC values from *GigaHeart* and competing methods in terms of linear probing on the CT-RATE dataset. For fine-tuning tasks (**A, B, C, E**), we used a subset (3,040 CTs) from CT-RATE to manage computational costs, while for linear probing (**D**), we leveraged the full dataset. In summary, *GigaHeart* outperforms competing methods substantially on almost all cardiovascular/thorax-relevant benchmarks in the aforementioned categories of tasks.

To ensure that the superior performance of *GigaHeart* is generalizable, we undertook cross-device evaluations. Here, *GigaHeart* was fine-tuned on CT scans produced from Siemens (Munich, Germany) machines and then tested on CTs from GE (Boston, USA) scanners (and vice versa). We observe that *GigaHeart* exhibits a strong cross-device generalization on five cardiovascular relevant abnormalities (Fig. 2B), attaining 8.9% AUROC and 9.1% AUPRC improvements on average over ResNet-50.

Next, we further assess the generalizability of our model on a cross-cohort setting. Specifically, we utilized two cohorts, one compiled by Duke [24] and another from IMU Mega Hospital [17] (Fig. 2C**, Supplementary** Fig. 5). *GigaHeart* demonstrated marked improvements in cardiomegaly, atelectasis and pericardial effusion detection. In this setting, *GigaHeart* shows a large improvement of 42.4% AUROC compared to the competing approach.

Linear probing, which freezes model parameters and only trains a linear classifier for downstream applications, is a more efficient approach to apply foundation models to real-world applications. We exploited linear probing to examine the quality of volume-level embeddings generated by different models and observed that *GigaHeart* achieves significant improvements on all tasks, confirming the high quality of embeddings from our model (Fig. 2D**, Supplementary** Fig. 6).

Finally, we exploited our organ-specific framework to develop a thorax-specific foundation model by focusing on thorax-region views. Similar to the promising performance on cardiac-specific tasks, this thorax-specific foundation model achieves the best performance on 9 thorax-related tasks (Fig. 2E**, Supplementary** Fig. 4). Similarly, ablation experiments show the thorax-aware training significantly improve the performance compared to standard DINOv2 training. (Supplementary Fig. 2) These results suggest the versatility of our framework, underscoring its potential to build organ-specific foundation models from CT data.

### *GigaHeart* enables multimodal cardiology applications

After confirming the superior performance of *GigaHeart* on unimodal tasks, we next examined how *GigaHeart* fares on multimodal tasks by joint modeling CT images and radiology reports. First, we applied vision-language alignment to *GigaHeart* by embedding together each CT volume and its corresponding radiology report in a new space (Fig. 3A). To do so, we finetuned *GigaHeart* using CLIP on 47,138 CT volume-radiology report pairs from CT-RATE. *GigaHeart* is used as the vision encoder and ClinicalBERT is used as the language encoder. The CLIP alignment endowed *GigaHeart* with multimodal understanding and zero-shot classification capabilities. *GigaHeart* demonstrates significant improvements on all three cardiovascular and six thoracic-relevant tasks over CT-CLIP (Fig. 3C, D**, Supplementary** Fig. 7**, 8**). Ablation studies replacing the language encoder with PubMedBERT showed a clear decline in performance. To further examine the quality of cross-modal alignment, we next evaluated two cross-modal retrieval tasks: identifying the closest matching chest CT volume given a radiology report and retrieving the most clinically consistent report given a CT volume. *GigaHeart* achieved a 77.5% recall improvement on volume-to-report retrieval and a 56.0% recall improvement on report-to-volume retrieval over CT-CLIP(Fig. 3E).

**Figure 3:**
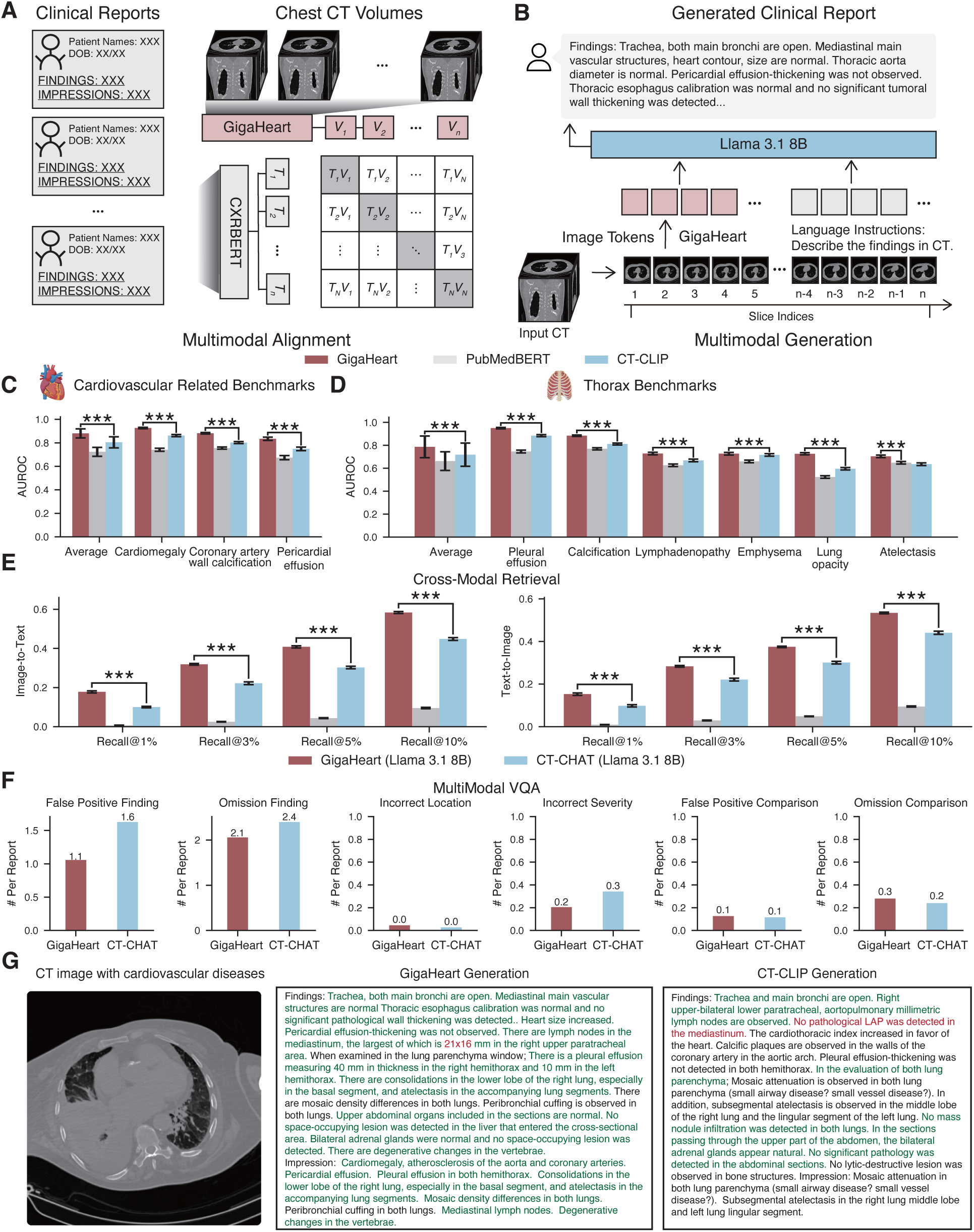
*GigaHeart* enables multimodal cardiology applications **A,B** Flow chart showing the fine-tuning of *GigaHeart* using radiology reports for multimodal alignment and generation. Multimodal alignment starts with training *GigaHeart* with a CLIP loss. Each CT volume and its associated report are considered positive pairs in contrastive loss. Then we proceed with a standard LLaVA training recipe, in which the inputs are CT volumes and language instructions. The outputs are responses, including answers to clinical meaningful questions or radiology reports. **C,D,** Bar plots comparing *GigaHeart* and CT-CLIP on cardiovascular and thorax related benchmarks on zero-shot tasks. **E,** Barplots comparing *GigaHeart* and competing methods on image-to-text and text-to-image retrieval. **F,** Barplots comparing *GigaHeart* and competing methods on report generation in terms of six clinically-relevant error types. **G,** A case study comparing the report generation of *GigaHeart* versus CT-CLIP. Green text indicates agreement with the ground truth report; red denotes disagreement. Overall, *GigaHeart*’s output shows stronger alignment with the ground truth. In all, *GigaHeart* outperforms CT-CLIP on zero-shot classification, cross-modal retrieval, multimodal VQA, and report generation tasks.

To enable multimodal generation, we adopted the LLaVA multimodal framework by using *GigaHeart* as the vision encoder and LLaMA-3.1-8B model as the language model (Fig. 3B). To ensure a comprehensive evaluation, we mainly considered four distinct types of visual question answering (VQA) tasks: (1) open question answering with short text, (2) open question answering with long text, (3) multiple choice answering, and (4) report generation. To evaluate the model’s responses, we reported both lexical similarity metrics, such as BLEU-1 [25], METEOR [26], ROUGE-L1 [27] and CIDEr [28], and LLM-based scoring metrics, which focus on clinical accuracy. *GigaHeart* consistently outperforms CT-CHAT, a multimodal AI assistant based on CT-CLIP, on all tasks across five metrics (Supplementary Fig. 9), underscoring its multimodal ability for jointly modeling CT images and reports.

Finally, we evaluated the factual accuracy of report generation in terms of six error types using CheXprompt [11]: false positives, omissions, incorrect location, incorrect severity, comparison false positives, and comparison omissions. The radiology reports generated by *GigaHeart*show 15.2% less errors on average (Fig. 3F). We have included a representative case to show that *GigaHeart* can generate a report that is more faithful to the ground truth report; *GigaHeart* successfully detects cardiothoracic abnormalities including cardiomegaly, atherosclerosis, pericardial effusion and pleural effusion, whereas these findings are not picked up by CT-CHAT (Fig. 3G). Taken together, these results highlight the effectiveness of *GigaHeart* as a more advanced multimodal AI assistant in real-world clinical tasks, such as generating highly accurate reports from chest CT image volumes.

### *GigaHeart* solves the oversizing problem in recipient-donor heart size matching by accurate segmenation

Heretofore, many of the tasks *GigaHeart* has been finetuned for (e.g. abnormality detection) are performed by humans (viz. radiologists) everyday as part of clinical care. To demonstrate the versatility of *GigaHeart*, we applied it to a clinical application for which the state-of-art practice typically involves neither AI nor chest CTs.

In heart transplantation, clinicians must determine whether a donor heart is too large (oversized) or small (undersized) for a given recipient. The current practice is to use a set of equations to calculate the predicted heart mass (PHM) for both the donor and recipient [6]. The equations are derived from regressing age, sex, age, and weight onto cardiac volumes segmented using a computer-assisted software program on cardiac MRIs of normal individuals [29, 30]. Donors and recipients who differ beyond certain thresholds (about 10%) have been found to have decreased survival after heart transplantation [5, 31].

Since chest CTS are almost always available for donors and recipients, it stands to reason that these images may provide more precise measures of cardiac size than PHM. Here, we use volume and mass interchangeably, because one can be converted to the other with a density factor. To avoid oversizing Fig. 4A, one simply needs to segment the volumes of the heart, i.e. derive the so-called total cardiac volumes (TCV) [32], from the chest CTs of both the donor and recipient. To do so, we finetuned *GigaHeart* on a dataset composed of segmented heart volumes from the TransUNet study [33] as well as 70 masks we hand-segmented.

**Figure 4:**
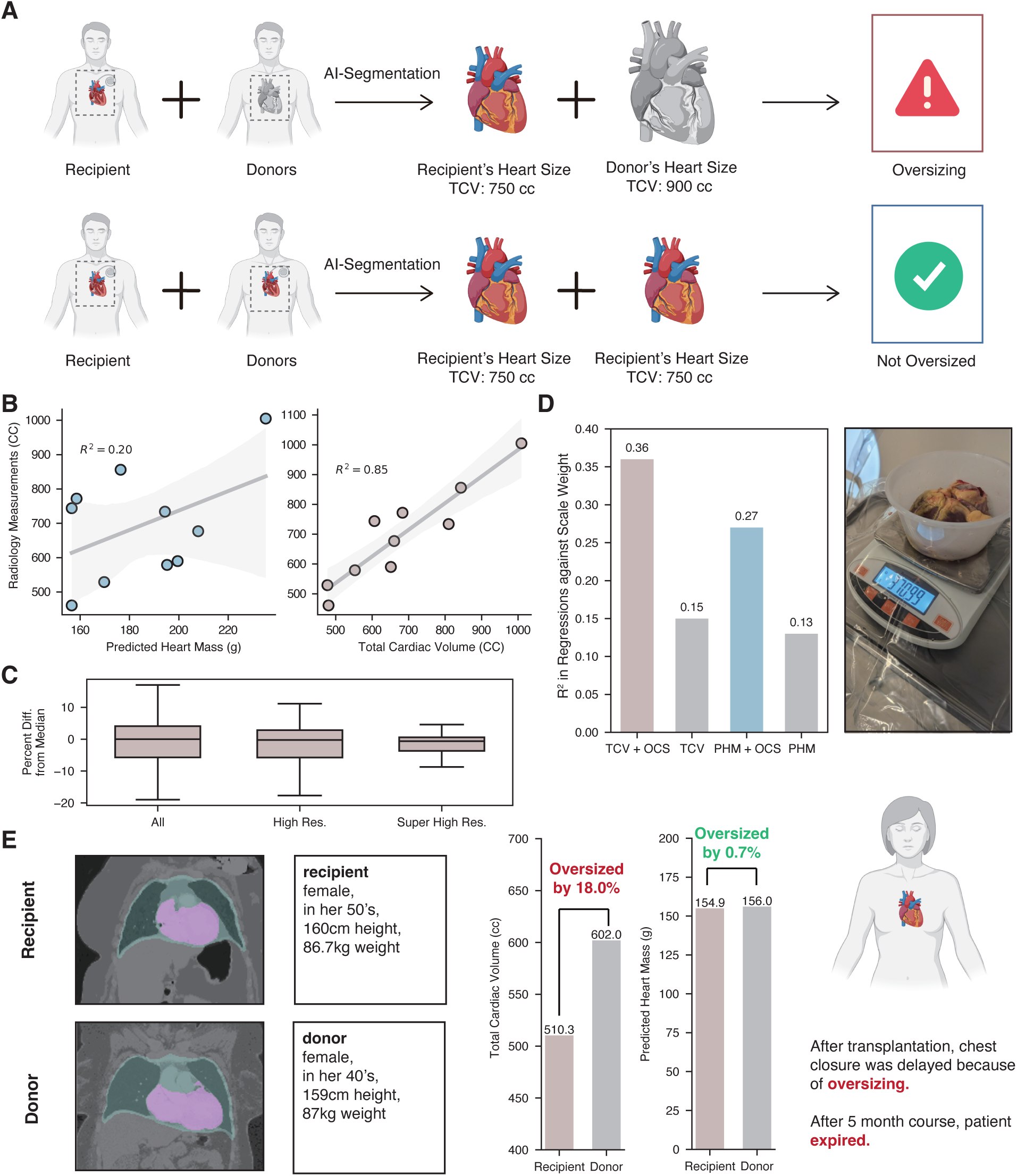
*GigaHeart* solves the oversizing problem in recipient-donor heart size matching by accurate segmenation. **A,** Flow chart illustrating the prevention of oversizing the donor heart for a recipient with *GigaHeart*. **B,** Regressions of data in the scatter plots show *GigaHeart* -estimated total cardiac volumes (TCV) are more accurate than PHM with radiologist-segmented volumes as the gold standard. **C,** Boxplots show the variation of estimated TCVs under varying resolutions. At the highest resolutions, the chest CT TCVs exhibit little variation irrespective of what types they are (e.g. contrast, non-contrast, non-gated, etc.). Thus, any type of chest CT can be used by *GigaHeart* for accurate segmentation, so long as it is higher resolution. **D,** Barplots comparing the regression R^2^ of TCV and PHM against heart mass obtained from weighing the heart on a scale in the operating room with or without the confounding variable OCS. When the confounder of whether the OCS system was used is included, TCV is shown to be more accurate than PHM in predicting the cardiac mass. **E,** Case study demonstrating successful prevention of donor heart oversizing. The PHM-generated values suggested the donor and recipient hearts were well-matched for size. However, the patient’s chest could not be closed at the end of the surgery, which led to infections and ultimately, the patient’s demise. In post-hoc analysis using *GigaHeart*, the precise measurements of the donor and recipient heart sizes from their personal imaging studies demonstrate that the donor heart was 18% larger than the recipient’s. Had this information been known, the heart would have been declined for transplant.

To compare our model with PHM, we first applied both methodologies to electrocardiogram (ECG)-gated CT angiograms of the heart. We gathered 10 CT angiograms of individuals who did not have left or right ventricular enlargement more severe than mild. To make the hearts more comparable with each other, we restricted images to be between 40% to 75% of the RR interval on their ECGs. For individuals who had several CT images within this interval, the TCVs were averaged. The gold standard used in this analysis were cardiac volumes derived from segmentations using radiologist-guided software. Fig. 4B demonstrates that the TCVs matched the radiologist-segmented volumes with much greater precision than PHM. The R^2^ for the regressions of TCV and PHM against the radiologist-segmented volumes were 0.85 and 0.20, respectively, with corresponding p-values of 0.0001 and 0.19.

In clinical practice, however, ECG-gated cardiac CT angios are not necessarily readily available for donors and recipients. More commonly, chest CTs of a great variety (different resolutions, contrast or non-contrast, mostly non-gated) will be given to clinicians deciding on the appropriateness of sizing between donors and recipients.

In order to give an idea of what the variance is for the different CTs of an individual, we take non-gated CTs of the same individuals and calculate the percent difference from the median of the various TCVs. Fig. 4C shows that the interquartile range is fairly modest, and outliers disappear as the TCVs of only higher resolution images are kept.

For further evidence that AI-segmented TCVs can be practically used, we again compared our AI model results with PHM using non-gated, donor CTs used in the heart tansplantation setting. We restricted the CTs examined to only high resolution ones. As the gold standard, we acquired the cardiac mass by weighing donor hearts on a scale just prior to implantation. It is widely known in the field that donor hearts which are placed on a normothermic organ preservation system (OCS, Transmedics, Andover, USA) become larger from edema, so this feature was included in the regression. Regressions of PHM and OCS against heart mass were found to have a higher R^2^ and lower p-value (R^2^ = 0.36, F-statistic p-value = 0.00003) when compared to the same analysis using PHM (R^2^ = 0.27, F-statistic p-value = 0.0005) (Fig. 4D).

With accurate TCVs of the donor and recipient, one can avoid a transplantation in which the donor heart is oversized for the recipient. Fig. 4E shows a real-life case study at our institution, in which a donor and recipient were well matched in their PHMs. After proceeding with transplantation, the donor heart was so large that the chest could not be closed during the index surgery. This complication led to difficult to treat infections and ultimately to the recipient’s demise 5 months after transplant. In retrospect with the benefit of applying *GigaHeart* to their chest CTs, we see that the donor heart was oversized by 18%, a fact that if it were known beforehand would have led to the surgery being called off. In this particular case, the recipient had restrictive cardiomyopathy and had a normal sized heart. Typically, recipients have enlarged hearts from their disease process. Having the TCV of an enlarged heart should expand the upper limit of acceptable heart sizes versus using PHM (Fig. 5E), so using this method should potentially lead to more donor heart acceptances.

**Figure 5:**
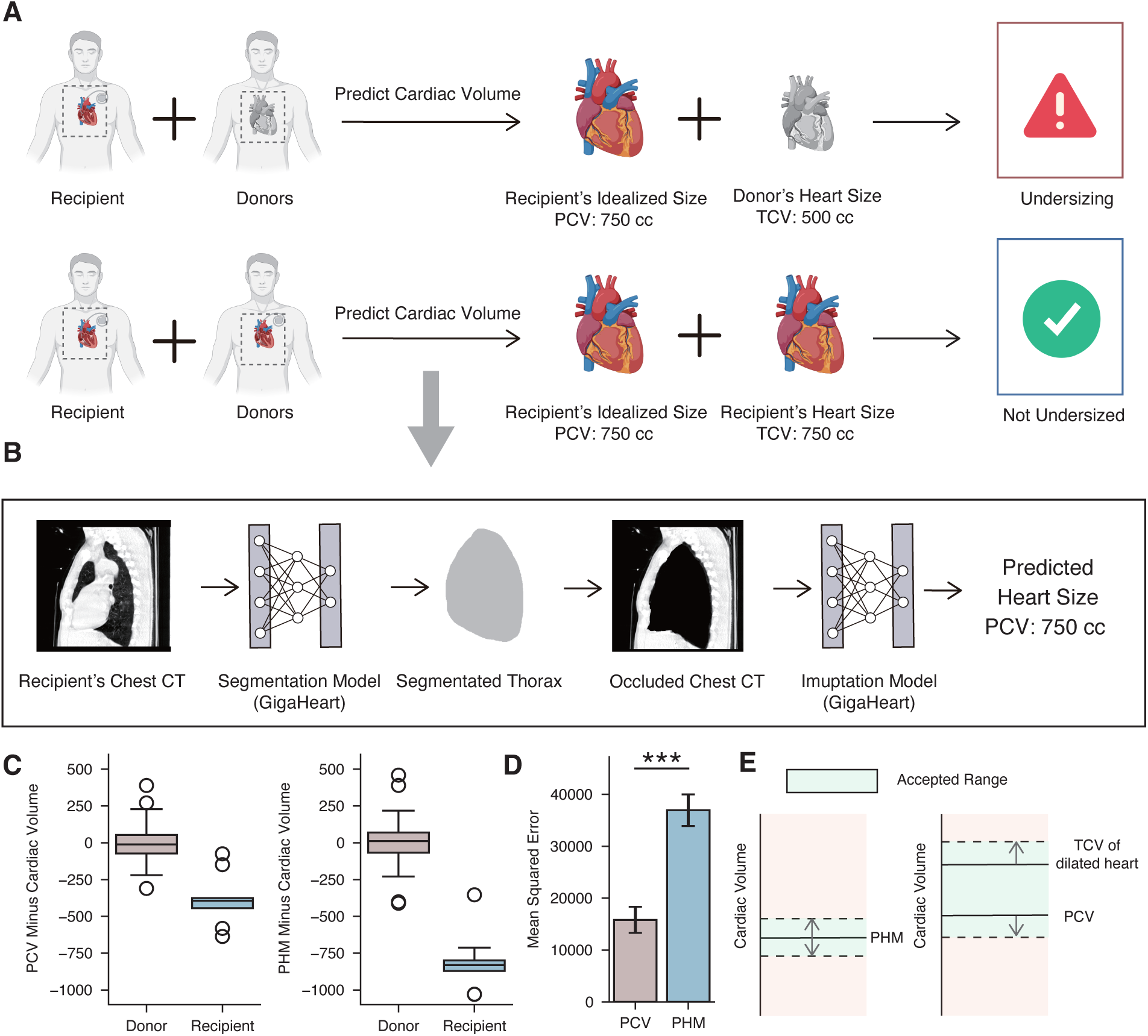
*GigaHeart* solves the undersizing problem in recipient-donor heart size matching through AI regression. **A,** Flow chart illustrating the prevention of undersizing the donor heart for the recipient with *GigaHeart*. **B,** Flow chart illustrating the pipeline of imputing a normal heart volume in a diseased recipient. The thoracic contents are first segmented and this volume is occluded in the original chest CT image. With this modified image, a heart volume (i.e. the predicted cardiac volume or PCV) is then imputed by using *GigaHeart* endowed with a regression layer. Since this model was trained on chest CTs with only normal-sized hearts, the regression output should represent a normal heart volume. **C,** Barplots comparing the mean squared error (MSE) of PCV and PHM. PCVs and PHMs are compared to the TCVs of chest CTs of individuals with normal-sized hearts. The MSE of the PCVs is lower than that of PHMs in a four-fold cross validation scheme. **D,** Boxplots illustrating differences in PCV vs. TCV and PHM vs. TCV among donors and recipients. Using previous regressions against TCVs, predicted values of the PCVs and PHMs are calculated and then subtracted from corresponding TCVs. Since donors have normal-sized hearts, the differences in these values should be near zero. For recipients, the differences should generally be negative, because the diseased hearts of recipients are oftentimes bigger than their idealized, normal heart sizes. We see these expected trends when using either the AI-generated PCVs or PHMs. **E,** Illustration of the concept of expanding acceptance ranges when using *GigaHeart* for heart sizing. When performing heart sizing with PHM, a clinican will make sure the PHM of the donor falls within about 10% above or below the PHM of the donor. If one uses *GigaHeart*, calculation of 10% above the TCV of the recipient as the upper bound, should in many cases, lead to a wider acceptable range, because the heart size of the diseased, recipient heart is oftentimes enlarged.

### *GigaHeart* solves the undersizing problem in recipient-donor heart size matching through AI regression

To avoid undersizing (Fig. 5A), however, the TCV of the recipient cannot be used as is, since, as previously mentioned, the diseased heart of the recipient prior to transplant is oftentimes dilated. What is needed is the idealized normal heart volume of the recipient if he/she had never developed heart failure; we term this volume the predicted cardiac volume (PCV).

To solve this problem, we implemented an AI regression model. We first trained a model to segment the thoracic contents (lungs, heart, mediastinum, etc.) from chest CTs. These volumes were then used to occlude those regions of the chest CT. We then trained a model to impute the size of a normal-sized heart (i.e. PCV) into the occluded space (Fig. 5B) via AI regression. Briefly, We took *GigaHeart* and added an additional layer for regression. This model was trained on 68,430 chest CTs of individuals with normal-sized hearts, whose cardiac volumes were ascertained with the previously developed TCV segmentation model. The rationale here is that since the this model has only seen normal-sized hearts, the cardiac volume it imputes for the occluded CT volume space should be normal-sized.

To validate our normal heart-size imputation model, we compare the PCVs to the PHMs using as the gold standard hand-segmented cardiac volumes of chest CTs previously employed to train the TCV segmentation model. For donors (i.e. patients with normal-sized hearts), we regress PCVs and PHMs separately against the hand-segmented cardiac volumes and calculate the MSE in a four-fold cross-validation scheme. We see that the MSE of the PCVs is lower than that of the PHMs (1.6 x 10^4^ and 3.7 x 10^4^ respectively, p = 1.2 x 10^−^6 by the Wilcoxin signed-rank test) (Fig. 5C).

We then perform regressions on all PCVs and PHMs separately against the hand-segmented cardiac volumes for donors only. We use the model coefficients to obtain the predicted cardiac volumes using the PCVs and PHMs for both donors and recipients. Boxplots (Fig. 5D) of these values minus the hand-segmented cardiac volumes show that as expected, the differences are close to zero for donors using either PCV or PHM as features. Also as expected, the differences are less than zero for recipients since the hand-segmented cardiac volumes of recipients oftentimes represent dilated hearts while the predicted cardiac volumes using PCVs or PHMs should be normal sized.

### *GigaHeart* accurately segments cardiothoracic anatomy and abnormalities

Finally, we examined whether *GigaHeart*, by focusing on the heart region, can also improve the segmentation of other anatomical structures and abnormalities. To cover diverse cardiothoracic segmentation tasks, we collected 122,910 chest CTs with segmentation masks on structures such as the heart, thorax, lung, esophagus, and spinal cord as well as CT findings such as lung nodules, lung tumors, and COVID-19 infection changes from five public datasets and proprietary data from UW Medicine (Fig. 6A). To benchmark *GigaHeart*, we chose SAM, MedSAM and SegVol as comparison approaches. Since SAM and MedSAM require a spatial prompt in addition to input images, we used a dummy bounding box as input and finetuned them on each dataset. The quantitative evaluation results demonstrated the superior performance of *GigaHeart*, surpassing SegVol by 84.9% and MedSAM by 48.7% on average in terms of the scores (Fig. 6B**, Supplementary** Fig. 10). Among these tasks, segmenting lung tumor, lung nodules, COVID-19 infection, esophagus and spinal cord is much more challenging due to the small object sizes and irregular shapes. Interestingly, *GigaHeart* achieved even greater performance gains, highlighting its superior sensitivity in capturing subtle anatomical and pathological details. For example, on lung nodule segmentation, *GigaHeart* achieved a Dice score of 0.6, while all comparison approaches have less than 0.2 Dice scores. Next, we considered using the oracle bounding box obtained from ground truth masks as inputs to SAM and MedSAM, *GigaHeart* still outperformed them on average by a large margin (Supplementary Fig. 11, Supplementary Fig. 12). Finally, we conducted qualitative analysis to compare *GigaHeart* segmentation results and ground truth. These examples show *GigaHeart* can accurately segment major organs like the heart and lungs, as well as detect subtle, irregularly-shaped structures, including COVID-19 lesions and small lung tumors.

**Figure 6:**
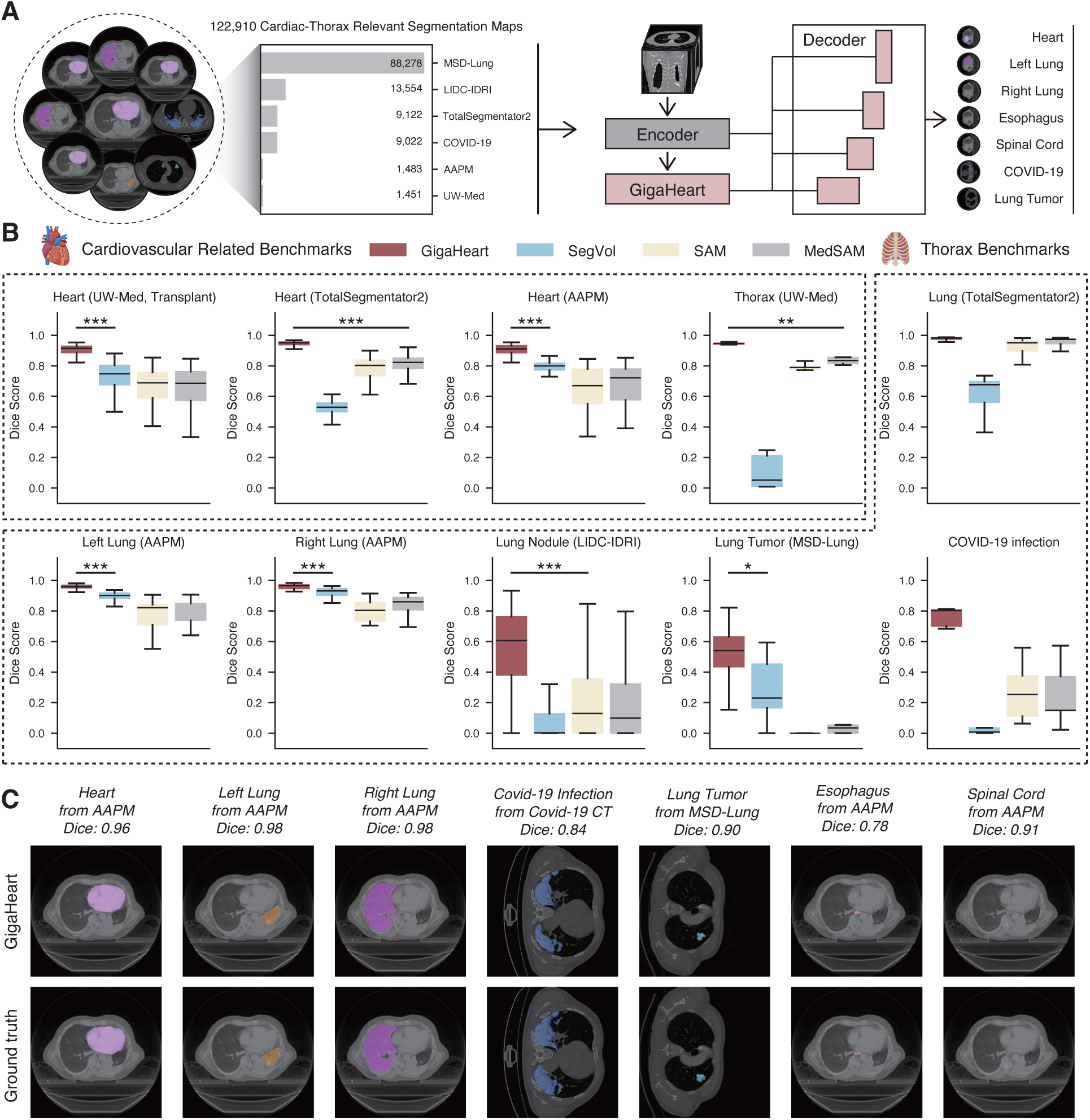
*GigaHeart* accurately segments cardiothoracic anatomy and abnormalities. **A,** Flow chart showing the adaptation of *GigaHeart* to a segmentation model. We collected 122,910 segmentation maps to cover a diverse set of cardiothoracic segmentation tasks. To adapt *GigaHeart* into a segmentation model, we follow the standard encoder-decoder structure used in TransUNet. **B,** Box plots comparing *GigaHeart* and competing methods on segmenting anatomy and abnormalities. Our model significantly outperforms SegVol, SAM and MedSAM on 10 out of 12 tasks from six datasets, including UW-Med, TotalSegmentator2, AAPM, LIDC-IDRI, MSD-Lung and Covid-19 Infection. **C,** Visual example demonstrating *GigaHeart*’s capability of segmenting cardiothoracic anatomic structures and abnormalities.

## Discussion

In this work, we show that *GigaHeart* performs better than competing models on a wide range of applications. We attribute its superior performance to several factors. First, in contradistinction to, for example, Merlin [18], which was trained on abdominal CTs, our pretrained model used chest CTs. Second, our pretraining set of chest CTs was, to our knowledge, the largest one to date; we used 180,897 chest CT volumes compared to 50,188 used by CT-CLIP [17]. Finally, our model learning incorporated the additional innovative step of heart-aware pretraining.

That *GigaHeart* excels at abnormality detection as well as report generation suggests that it may be of utility in the clinical setting. We do not propose that *GigaHeart* can take the place of a human reader wholesale. However, its relative accuracy, computational quickness, and imperviousness to fatigue may help a radiologist improve his/her accuracy and throughput by making preliminary findings that he/she can overread for finalization.

Beyond aiding in tasks which are widely performed by radiologists everyday, the foundation model paradigm can potentially facilitate the development of many more models for bespoke applications. The reason is that pretaining allows for better and perhaps even acceptably useful performance in the clinical setting of finetuned tasks when only a small set of labeled data can be practically acquired.

Recipient-donor heart-size matching is one such example of a niche application in which we have demonstrated that the finetuned application of our foundation model performs better than the state-of-art practice of using PHM. For the TCV segmentation task, it is not to say that methods do not currently exist for achieving the same end. Computer assisted software exists to accomplish this very task on chest CTs. However, it requires a radiologist (or radiology technician) to perform activities at times outside of normal working hours. The AI segmentation program offers an accurate, easy, and quick (a single segmentation takes minutes) alternative.

As the results of Fig. 4C demonstrate, our model is valid for heart size matching across a number of different types of chest CTs, irrespective of contrast usage, CT machine manufacturer, hospital of origin, etc. It is true that a dedicated, ECG-gated cardiac CT would yield even greater precision. However, obtaining these studies for most donors and recipients is not practical. They require (1) specialized radiology technicians for image acquisition; (2) the use of contrast agents, which can compromise kidney function and render these organs unsuitable for transplant; and (3) a low and stable heart rate, whereas donor and recipient heart rates are often elevated.

Even if heart volumes can be ascertained with perfect accuracy, obtaining these measures for the recipient and donor only solves the donor oversizing problem. We need to impute an idealized, normal heart volume of the recipient to ensure the donor heart is not undersized. For this task, admittedly, there is no practical gold standard which can be acquired. However, the use of PHM for potential heart transplant patients has become widely used by the heart transplant community solely based on its performance on normal individuals. In this work, we have shown that an AI regression model performs better than the PHM in heart donors (i.e. individuals with normal-sized hearts). The adoption of PCV instead of PHM on this basis should logically follow. If one takes a step back, perhaps it is not unintuitive that a method using the many bodily features outside the inner thoracic contents is more information-rich than a mere four factors to determine the idealized, normal size of a heart.

As stated in the results, since recipient hearts usually enlarge as part of the disease process, using TCVs of the donors and recipient should increase the upper limit of acceptable heart sizes than if PHM were used. Increases in acceptable donor hearts should lead to recipients getting transplants faster Fig. 5E. For the undersizing problem, using the AI regression method will not necessarily decrease the lower size limit as compared to PHM. However, more accurate imputation of a normal heart size for the recipient should lead to better donor-recipient matching from a size perspective and better transplant outcomes.

## Data Availability

Our model will be accessible at https://github.com/gigaheartmodel/gigaheart.git upon publication, including the model weights and relevant source code for heart size matching. We also include detailed methods in the Methods section. Our pretraining data will be accessible upon request.

## Acknowledgements

We would like to thank the Raisbeck family for providing unrestricted research funding for this project. We also acknowledge D. Y. Lin and W. Lin’s contributions to the segmentations of CT images.

## Methods

### Datasets used

From the University of Washington Medical Center echocardiography database, the medical record numbers of patients who had corresponding reports with normal left and right ventricular sizes from 01/01/2014 to 11/14/2023 were collected (set 1). Another set of medical record numbers were collected if the echocardiogram reports contained left ventricular enlargement under the same time period (set 2). Finally, we collected a third set of medical record numbers of individuals who had undergone heart transplant (set 3). Donor chest CTs were initially acquired from CompuMed (Los Angeles, USA) but later could be directly downloaded from the United Network for Organ Sharing (UNOS) website (set 4). The data were gathered under the approval of the University of Washington institutional review board.

Once these medical record numbers were collected, their corresponding chest CTs in digital imaging and communications in medicine (DICOM) format were compiled. These files were then resampled into 512 x 512 x 512 isotropic image volumes in which each pixel represented a cubic centimeter by using imaging software MIM (Cleveland, USA).

To arrive at a set of CTs whose individuals had normal-sized hearts used for training the AI regression model for imputing normal sizes (explained further below), sets 2 and 3 were subtracted from set 1. The reason is that an individuals may develop an enlarged heart over time, or their heart may have normalized after transplantation. The chest CTs corresponding to the remaining medical record numbers after set subtractions were considered to have normal-sized heart ventricles (set 5). Gated CT angios were chosen as follows. The number of chest CTs corresponding to each medical record number in set 5 were tallied and ranked. The top 10 with the most chest CTs, one of which had to be an electrocardiogram (ECG)-gated study, were chosen for further processing. The phases (position in the RR cycle) available for each study varied.

### Pretraining *GigaHeart*

Our pretraining method consisted of two-stages, the first of which involved DINOv2 [19] followed by organ-aware pretraining. For the first stage, we preprocessed 180,897 CT volumes into 60,320,854 2D image slices by applying min-max normalization on the raw values and then histogram equalization. We chose a 24-layer vision transformer with 1024 embedding dimensions and 16 attention heads, amounting to 300 million learnable parameters. The vision transformer resized the image into 256 × 256 pixels and tokenized the image into 16 × 16-pixel patches. We used a local batch size of 128 to train the vision transformer for 1,000 epochs on 4 A100-80GB GPU devices, leading to a global batch size of 512 and 1,250,000 iterations in total. We followed the default configuration in the standard DINOv2 training to setup the DINO head, the KoLeo loss weight, the iBOT loss, and the learning rate scheduler. We also used centering instead of the Sinkhorn-Knopp [19] centering in the teacher model.

For the organ-aware training of the second stage, our goal was to guide the model’s attention to specific organs of interest. We used the first-stage pretrained model as the backbone and adapted it as a segmentation model for specific organs with human segmented data. We trained the model on 70 CT volumes with expert annotations on the heart and thorax. Our approach used the very simple yet effective technique of teaching the model the location and size of organs of interest to further enhance its performance in downstream tasks. We only replaced the random multi-cropping in the initial DINOv2 pretraining with the organ-aware cropping strategy. In the organ-aware cropping, we iteratively apply random multi-cropping until the crop window either fully encloses the organ mask or is entirely contained within it. Finally, we continued the DINOv2 training with the organ-aware cropping for another 300 epochs, with the same configurations of local batch size, learning rate, and model architecture as in the first-stage pretraining. Using heart-aware and thorax-aware cropping, we produced the cardiothoracic foundation model tailored to the cardiac and thoracic regions.

### Gold Standards Used

To finetune *GigaHeart* to segment the heart, the training and test sets came from various sources. The first were chest CT images and volume masks from public datasets. We also included chest CT and heart volume masks (i.e. the total cardiac mass or TCV) from heart transplant recipients (set 3) and donors (set 4). These masks were from serial hand segmentations of 2D CT images performed using the MIM software (Cleveland, USA) and took about 4 hours each to complete. Though MIM has its own segmentation functions, substantial correction by a human was required. Thoracic masks (used for the imputing normal heart sizes) were also similarly hand-segmented and took about 8 hours to complete for each series.

For the ECG-gated CT angiograms, the radiologist-calculated TCV was found with the assistance from the radiographic software TeraRecon iNtuition (TeraRecon Inc., Durham, USA). It took a trained radiologist approximately 15 minutes to attain the cardiac volume for each series at a single point in the RR interval using this program.

For the boxplots showing variance of studies over different resolutions Fig. 4D, the operational definition of high resolution is that the study has 1) more than 100 slices and 2) the slice thickness is less than equal to 1.25 cm; for super high resolution, 1) more than 100 slices and 2) the slice thickness is less than 1.25 cm.

The heart masses of donors were acquired by weighing hearts on a scale lined by a sterile cover in the operating room. Prior to weighing, hearts were prepared for implantation by separating the aorta and pulmonary artery and cutting to length, closing a patent foreman ovale (if present), and shaping the left atrial cuff. After weighing, the heart was immediately implanted into the recipient.

### Details of finetuning *GigaHeart* on classification tasks

When finetuning the model on downstream tasks, we applied an additional one layer MLP-based classifier to the embeddings generated by *GigaHeart*. Specifically, for each input CT volume, we resized the number of slices, height, and width into 256 × 256 × 256 pixels while keeping its original aspect ratio. Then we uniformly downsampled the number of slices for each volume into 64 frames. Each frame is further converted into a 1024-dimension embedding vector and we average all 64 embedding vectors into a single volume-level embedding. The task-specific MLP classifier is then applied on top of this embedding to produce the predicted probability for each class. For each task, we finetuned both the parameters of *GigaHeart* and the classifier using a base learning rate of 2 × 10^−3^ with one CT volume in each mini-batch. We set the number of finetuning epochs to 5 with a weight decay of 0.05 and layer-wise decay of 0.95. We use the first epoch as the warm-up epoch. We use cross-entropy loss to train the model for both binary and multi-class classification tasks:

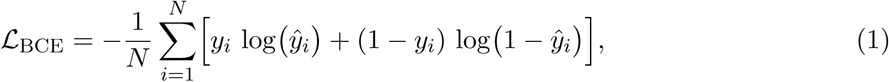

where *y_i_* is the ground truth label and *y*^*_i_* is the model prediction. For all the downstream tasks, we trained our model on a single A100 GPU device.

### Details of finetuning *GigaHeart* for multimodal understanding and generation

We first perform CLIP training to equip *GigaHeart* with multimodal understanding capabilities. We used the pretrained *GigaHeart* as the vision encoder and ClinicalBERT or BiomedBERT as the language encoder. We performed contrastive learning to push the image and their paired text embeddings to be close in the latent space. The contrastive learning loss is as follows:

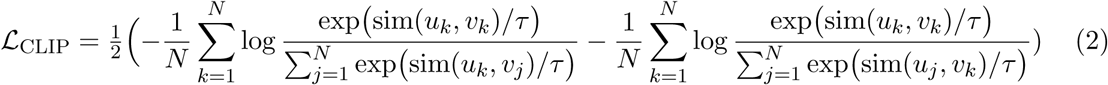

We use *u_k_* and *v_k_* as the CT image and text embeddings and *τ* as the temperature. To train the CLIP model, we used 8 80GB A100 GPU devices with a local batch size of 4 on each device. We also set the gradient accumulation steps to 32, which amounts to a global batch size of 1,024 in total. We set the training epochs to 5.

After CLIP training, we adapted our model for multimodal generation by combining *GigaHeart* and a language decoder using the LLaVA architecture. Specifically, we used the Llama 3.1 8B model as the language decoder. The input of the language decoder is the text tokens of user’s questions and the visual tokens of CT images. We used the multimodal VQA dataset in CT-RATE as training data, which covers four types: long answer, short answer, multiple choice and report generation. Following LLaVA, we first finetuned the MLP projector for 1 epoch using a learning rate of 1 × 10^−3^ with a batch size of 12. Then we finetune the language decoder for 3 epochs using a batch size of 3.

### Details of finetuning *GigaHeart* for normal heart size imputation via AI regression

We trained the normal heart-size imputation model on 68,430 normal heart CTs from 68,430 patients. In this study, among the multiple chest CTs a single patient can have, we pick the highest resolution ones for training this model. After the resolution-based selection, we ran our segmentation model to segment the thorax and heart. The input of the model is the chest CT for which we occlude the thorax using the AI-segmented thorax masks. The total cardiac volume is calculated using the sum over pixels across all slices, and this value is used as the regression target. We follow the same approach as the classification tasks to generate the volume-level embedding and use a shallow one-layer MLP to predict the cardiac volume. We trained normal heart-size imputation model using a base learning rate of 2 × 10^−4^ and batch size of 1 for 5 epochs. We use a weight decay of 0.05 and layer-wise decay of 0.95. We still use the first epoch as the warm-up epoch. We used the mean squared error (MSE) loss for training this model:

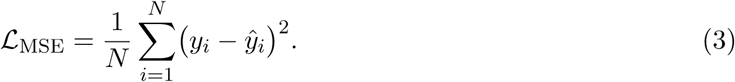

### Details of finetuning *GigaHeart* for segmentation

To adapt *GigaHeart* for segmentation tasks, decoding image representations in latent space into pixel space is necessary. Here we combine the encoder-decoder architecture from TransUNet and *GigaHeart* to be the ViT backbone of our segmentation architecture. We perform full finetuning to achieve strong performance on a wide range of downstream tasks. For pretraining, We collected a diverse set of segmentation data which consists of 122,910 cardiac-thorax relevant segmentation maps from five public sources as well as one in-house expert-annotated dataset from UW Medicine, covering 10 anatomy structures and disease segmentation tasks. To train the segmentation model, we chose the combination of the cross-entropy loss and Dice loss. The Dice loss can be written as:

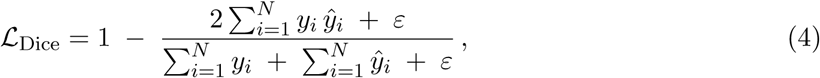

where the *y_i_* ∈ {0, 1} represents the ground truth mask at pixel i, and *y*^*_i_* is the prediction. The combined loss used for training is:

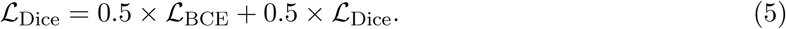

In our training, we use a learning rate of 2 × 10^−4^ with a batch size of 8. We set the number of training epochs to 20. The model is trained on a single 80GB A100 GPU.

### Details of comparison approaches

On image-based classification tasks, we considered Merlin, SegVol, ResNet-50 and the public DINOv2 checkpoint as the competing method.

1. **Merlin**, we obtained the model from: https://huggingface.co/stanfordmimi/Merlin. We initialized the model with the image embeddings active and obtained the embeddings which we use for downstream evaluation.
2. **SegVol**, even though with its main focus on segmentation tasks, it has a 3D ViT backbone and the ViT backbone was pretrained using SimMIM [34]. Here we investigated whether this pretrained 3D model can generate high quality image embeddings. We take the 3D ViT backbone out and applied a one-layer MLP as the classifier on downstream tasks. We obtained the model from the huggingface link: https://huggingface.co/BAAI/SegVol.
3. **ResNet-50**, we obtained this model from https://huggingface.co/timm/resnet50.a1 in1k. We adapted the ResNet-50 into a 3D version using the python library timm-3d, which replicates each 2D convolution kernel in ResNet-50 into 3D convolution layers. We applied an additional MLP layer on top of the 3D convolutional neural networks as the classifier.
4. **Public DINOv2**, here we used the public DINOv2 checkpoint out of box to generate an image embedding vector for each CT slice. The volume-level embedding is generated by mean pooling across slices. We obtained the public DINOv2 checkpoint from the link: https://huggingface.co/timm/vit large patch14 dinov2.lvd142m. To ensure a fair comparison, we use the same finetuning hyperparameters, with a base learning rate of 2 × 10^−3^ and a batch size of 1. We set the weight decay to 0.05 and the training epochs to 5.

On the multimodal zero-shot classification, cross-modal retrieval, the VQA, and report generation tasks, we chose CT-CLIP and CT-CHAT as competing methods. We directly use these pretrained models and compare our model against them on the CT-RATE dataset. Both CT-CLIP and CT-CHAT use the CT-RATE dataset to pretrain their model and we used the images not included in their training from the released dataset for evaluation purpose. We obtained the dataset and model from the link https://huggingface.co/datasets/ibrahimhamamci/CT-RATE/tree/main.

On the segmentation tasks, we considered SAM, MedSAM and SegVol as competing methods. Both SAM (SAM-ViT-B, initialized from https://huggingface.co/facebook/sam-vit-base) and MedSAM (MedSAM-ViT-B, initialized from https://huggingface.co/wanglab/medsam-vit-base) were fine-tuned for 50 epochs on 1 NVIDIA A100 GPU in mixed precision (FP16) with AdamW. Each axial CT slice was resized to 1024 × 1024 pixels. We set the learning rate to 1 × *e*^−5^ with five warmup epochs. SegVol was evaluated with the public checkpoint (https://huggingface.co/BAAI/SegVol) and tokenizer. Whole-volume CT scans are passed through SegVol’s zoom-in/zoom-out inference pipeline. Each test volume received a ground-truth bounding-box prompt and a class-specific textual prompt for the corresponding class (esophagus, heart, left lung, right lung, spinal cord, COVID-19 infection, lung nodule, lung tumor, lung, thorax); inference was performed on one A100 GPU.

## Statistical analysis

In comparing the performance of *GigaHeart* finetuned to specific tasks, the student’s t-test [35] implemented in package scikit-learn within programming language Python [36] was used. Regressions were performed using package scikit-learn. Mean squared error (MSE) comparisons between predicted cardiac volumes (PCV) and the predicted heart mass (PHM) in a four-fold cross validation scheme was performed with the Wilcoxin signed-rank test[37] implemented in the statistical programming language R [38].

## Data and Code Availability

*GigaHeart* will be accessible at https://github.com/gigaheartmodel/gigaheart.git upon publication, including the model weights and relevant source code for heart size matching. We also include detailed methods in the Methods section. Our pretraining data will be accessible upon request.

## Supplementary Information

**Supplementary Figure 1:**
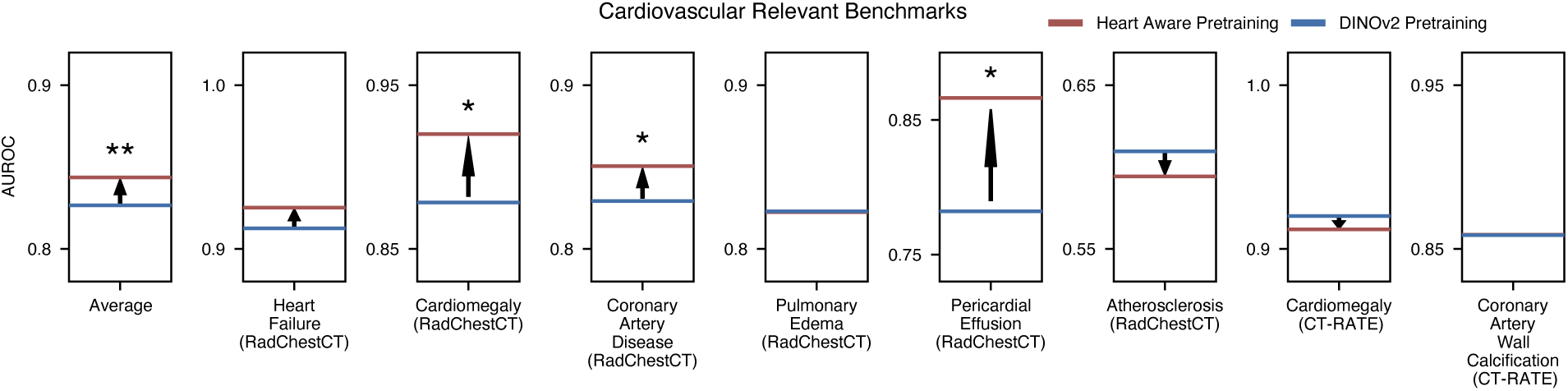
Ablation studies showing improvement of heart aware pretraining The statistical significance was determined using five-fold cross-validation.

**Supplementary Figure 2:**
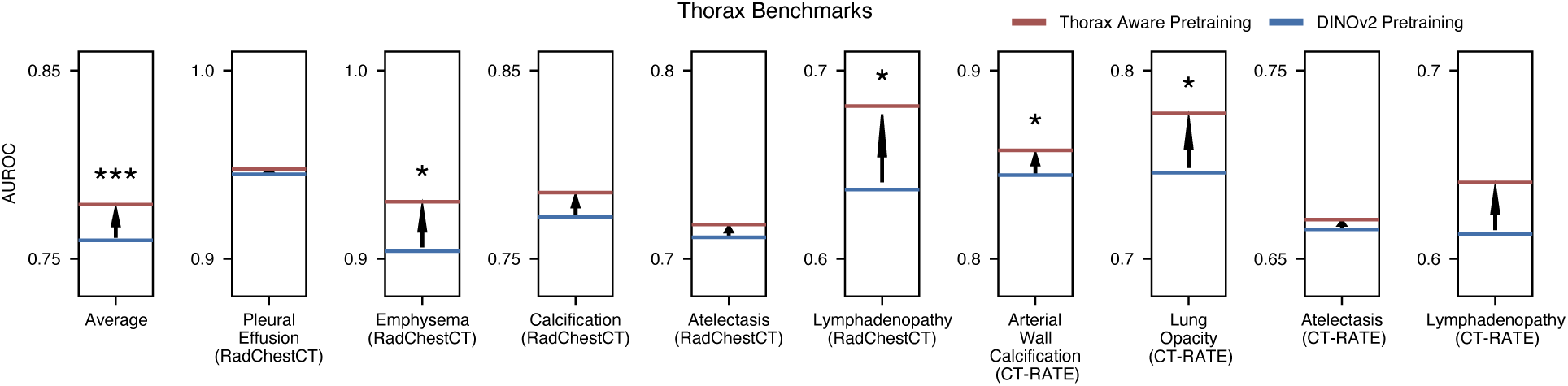
Ablation studies showing improvement of thorax aware pretraining The statistical significance was determined using five-fold cross-validation.

**Supplementary Figure 3:**
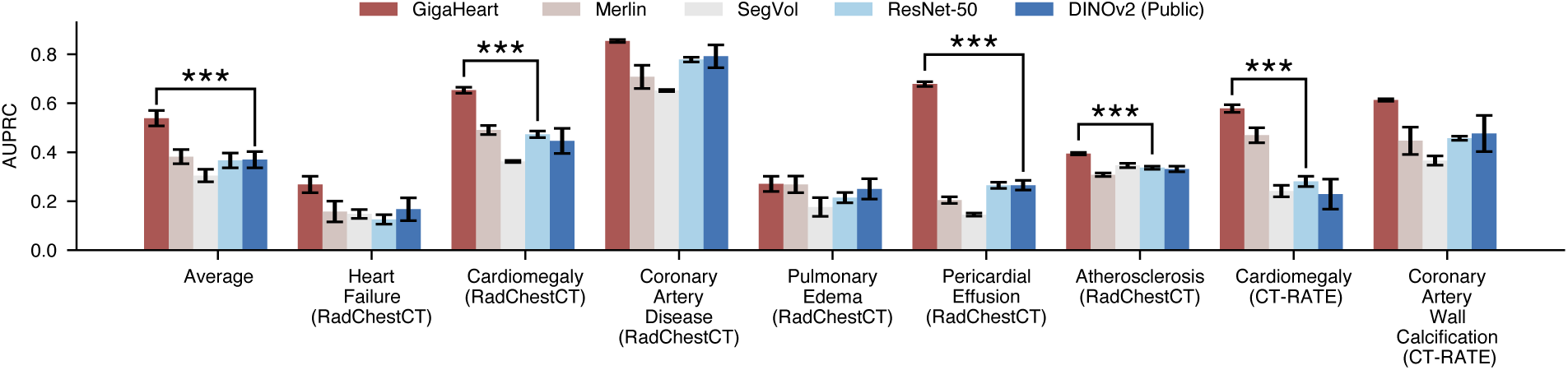
Bar plot comparing *GigaHeart* with competing methods on cardiovascular related benchmarks using AUPRC. ∗ indicates the significance level at which *GigaHeart* outperforms the best-competing method, with Wilcoxon test *p*-value< 1 × 10*^−^*^3^ for ***.

**Supplementary Figure 4:**
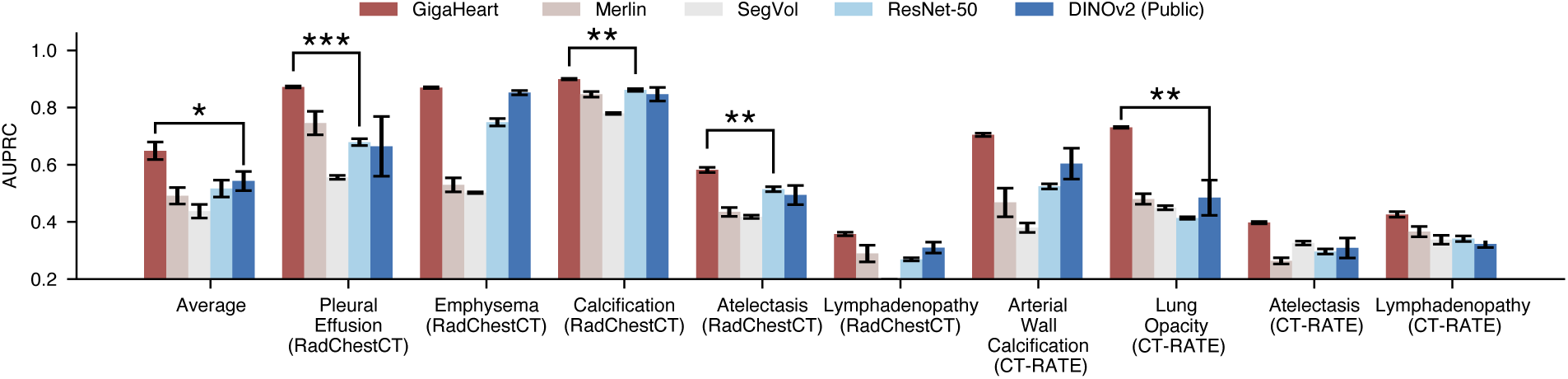
Bar plot comparing *GigaHeart* with competing methods on thorax benchmarks using AUPRC. ∗ indicates the significance level at which *GigaHeart* outperforms the best-competing method, with Wilcoxon test *p*-value< 5 × 10*^−^*^2^ for *, *p*-value< 1 × 10*^−^*^2^ for **, *p*-value< 1 × 10*^−^*^3^ for ***.

**Supplementary Figure 5:**
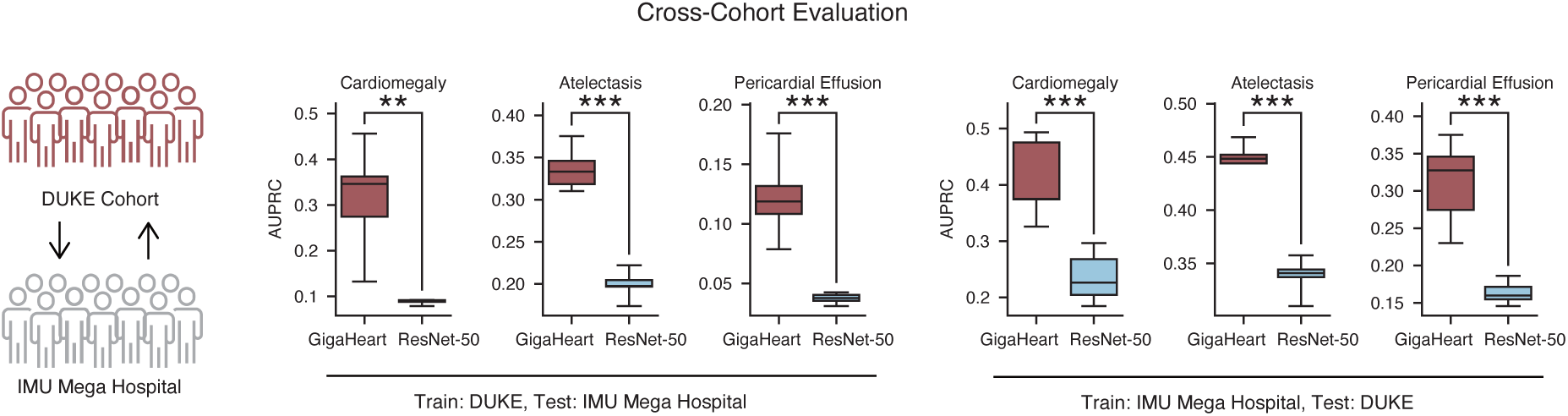
Bar plot comparing *GigaHeart* with ResNet-50 on a cross-cohort setting using AUPRC. ∗ indicates the significance level at which *GigaHeart* outperforms the best-competing method, with Wilcoxon test *p*-value< 1 × 10*^−^*^2^ for **, *p*-value< 1 × 10*^−^*^3^ for ***.

**Supplementary Figure 6:**
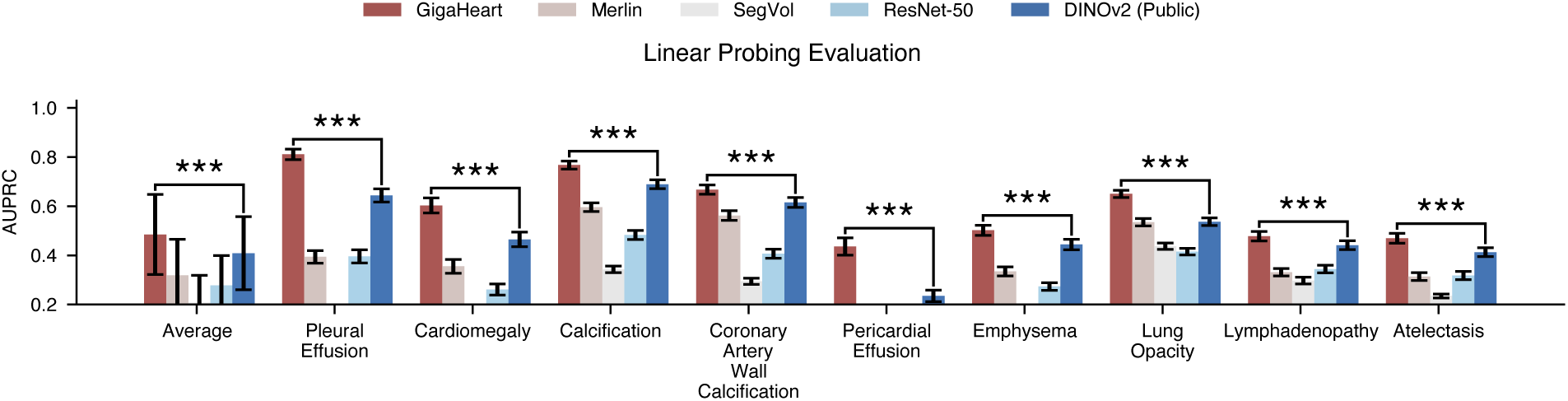
Bar plot comparing *GigaHeart* with competing methods on the linear probing setting using AUPRC. ∗ indicates the significance level at which *GigaHeart* outperforms the best-competing method, with Wilcoxon test *p*-value< 1 × 10*^−^*^3^ for ***.

**Supplementary Figure 7:**
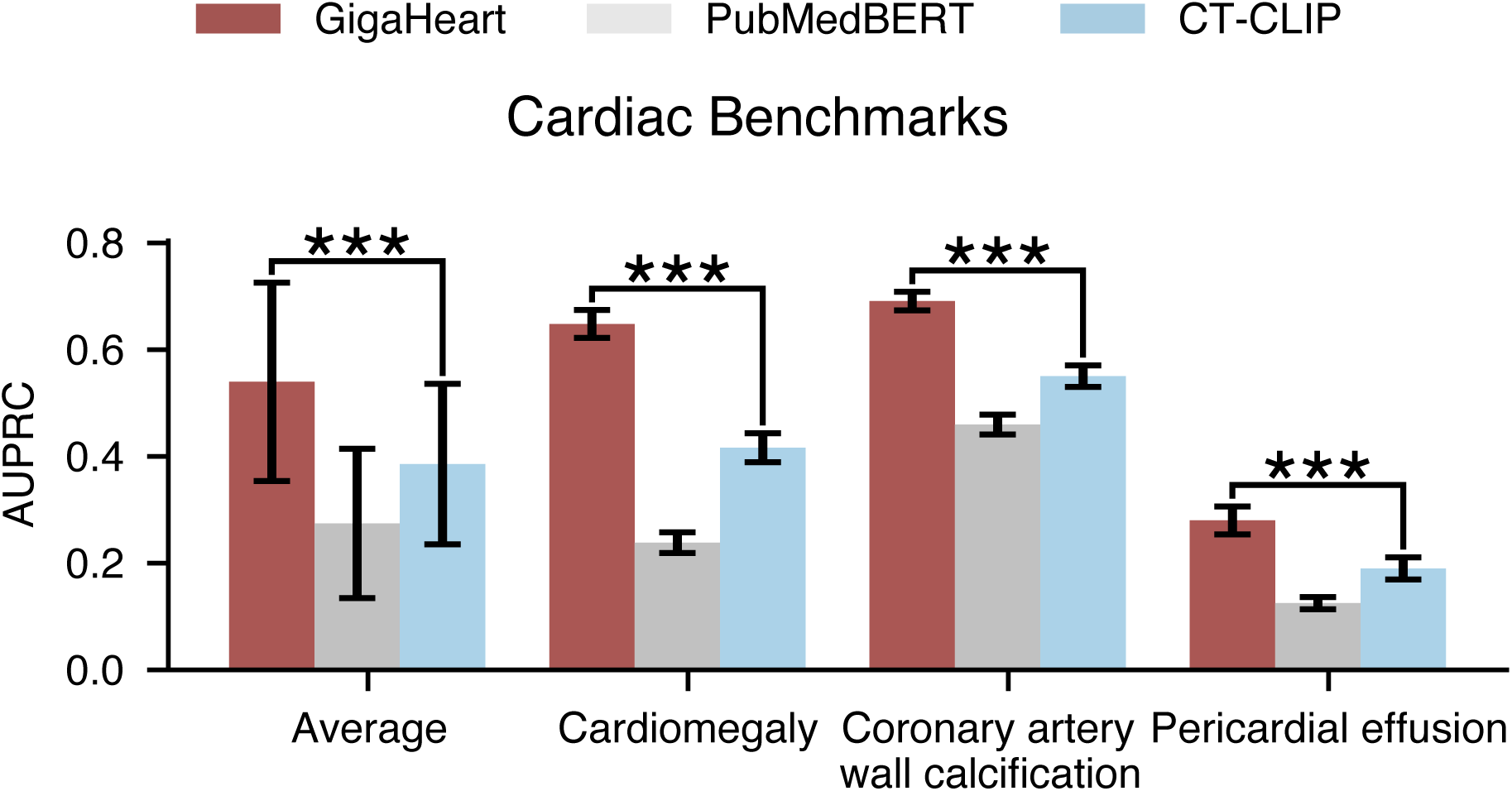
Bar plot comparing *GigaHeart* with competing methods on the zero-shot cardiovascular related benchmarks setting using AUPRC. ∗ indicates the significance level at which *GigaHeart* outperforms the best-competing method, with Wilcoxon test *p*-value< 1 × 10*^−^*^3^ for ***.

**Supplementary Figure 8:**
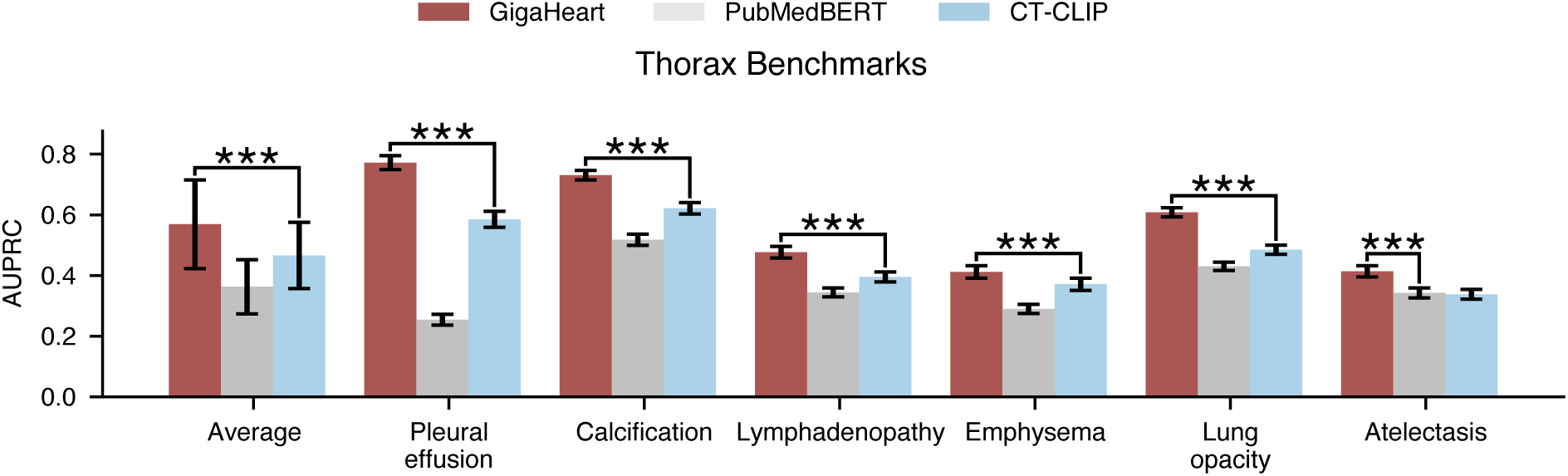
Bar plot comparing *GigaHeart* with competing methods on the zero-shot thorax benchmarks setting using AUPRC. ∗ indicates the significance level at which *GigaHeart* outperforms the best-competing method, with Wilcoxon test *p*-value< 1 × 10*^−^*^3^ for ***.

**Supplementary Figure 9:**
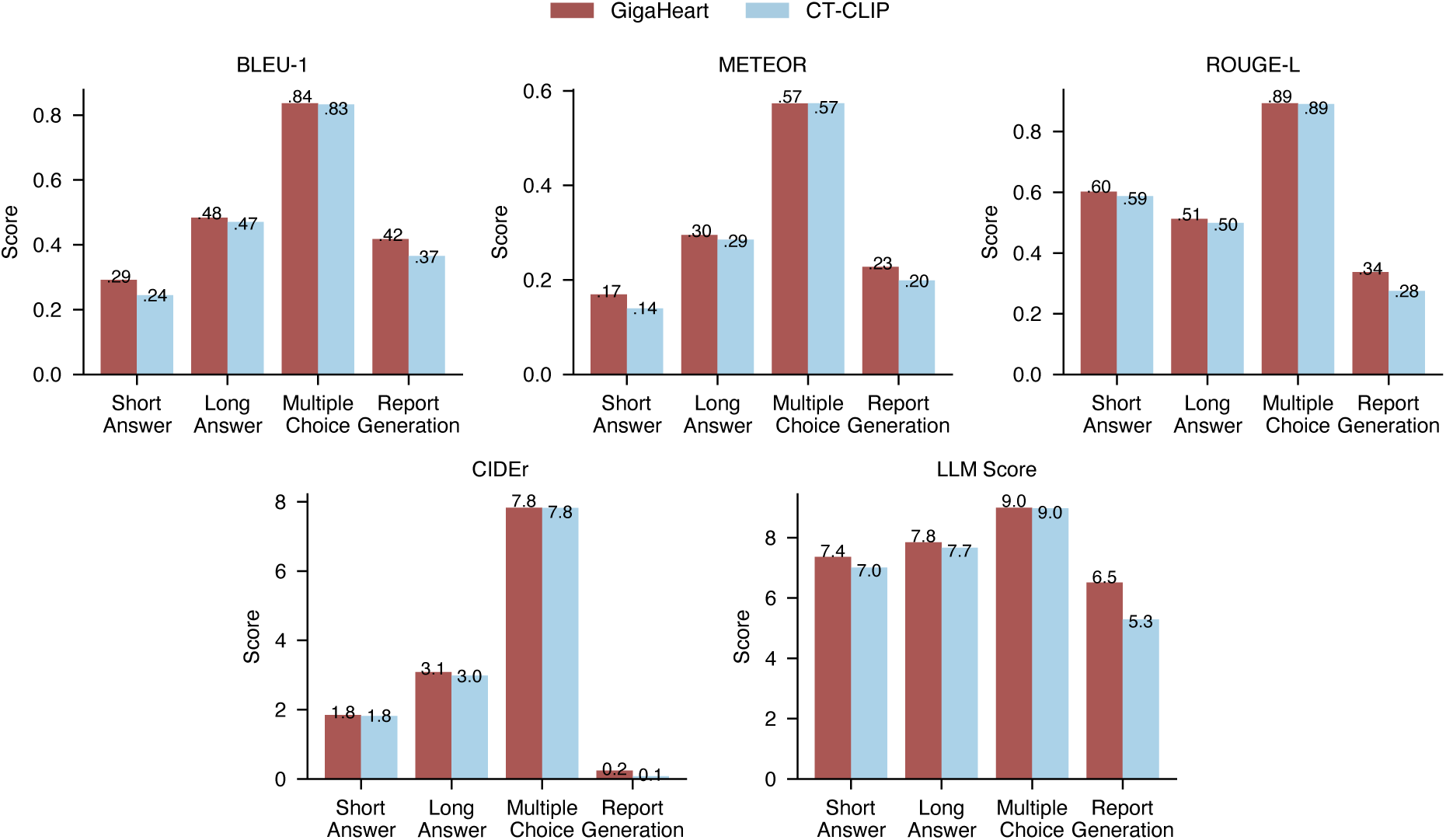
Bar plot comparing *GigaHeart* with CT-CLIP on the CT-RATE VQA and report generation benchmark. We compare *GigaHeart* with CT-CLIP using BLEU-1, METEOR, ROUGE-L, CIDEr and LLM score. LLM score is calculated with LLaMA-3.1 70B model. We followed the prompts used in CT-CLIP.

**Supplementary Figure 10:**
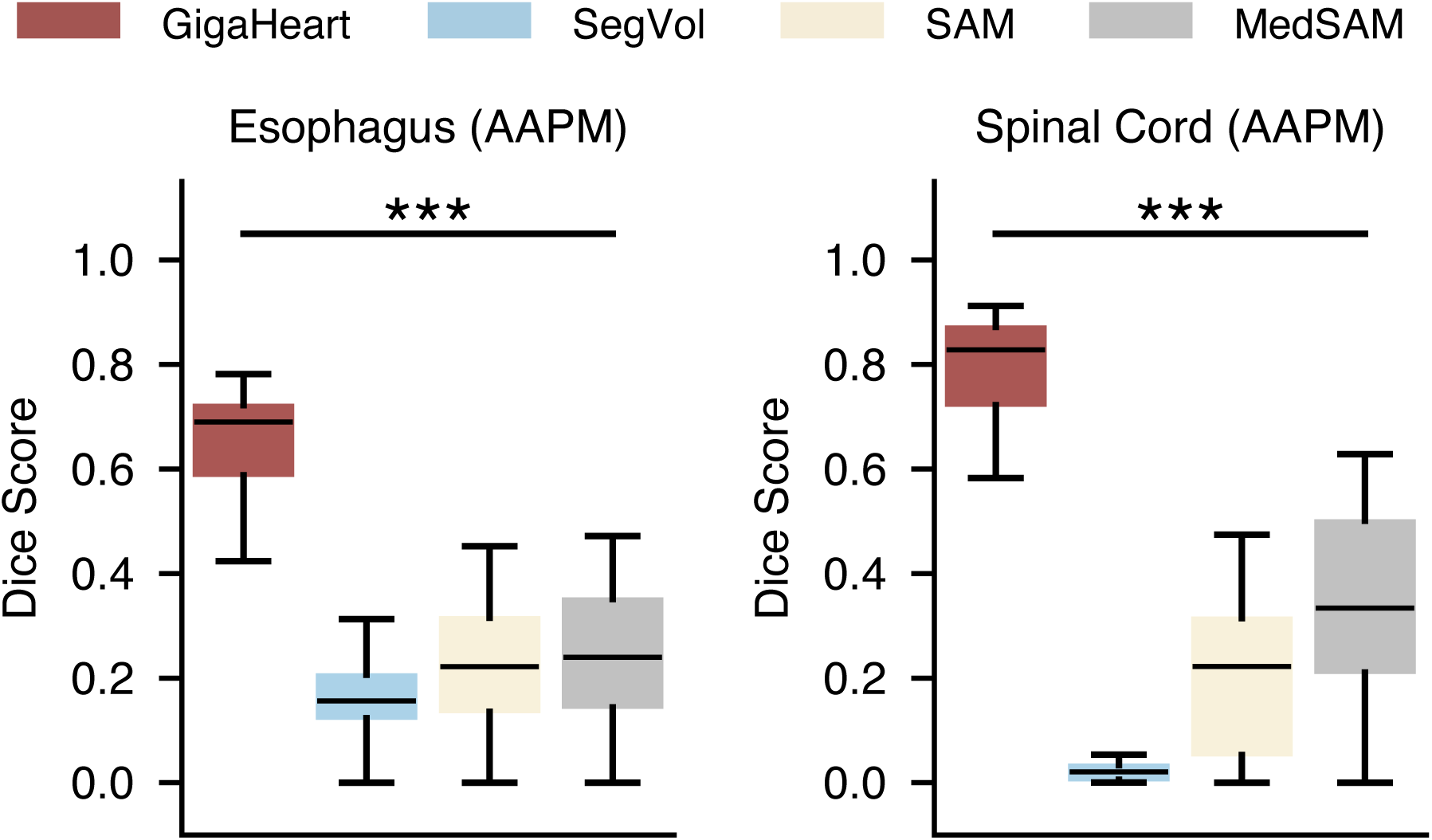
Box plot comparing *GigaHeart* with competing methods on segmenting the esophagus and spinal cord. ∗ indicates the significance level at which *GigaHeart* outperforms the best-competing method, with Wilcoxon test *p*-value< 1 × 10*^−^*^3^ for ***.

**Supplementary Figure 11:**
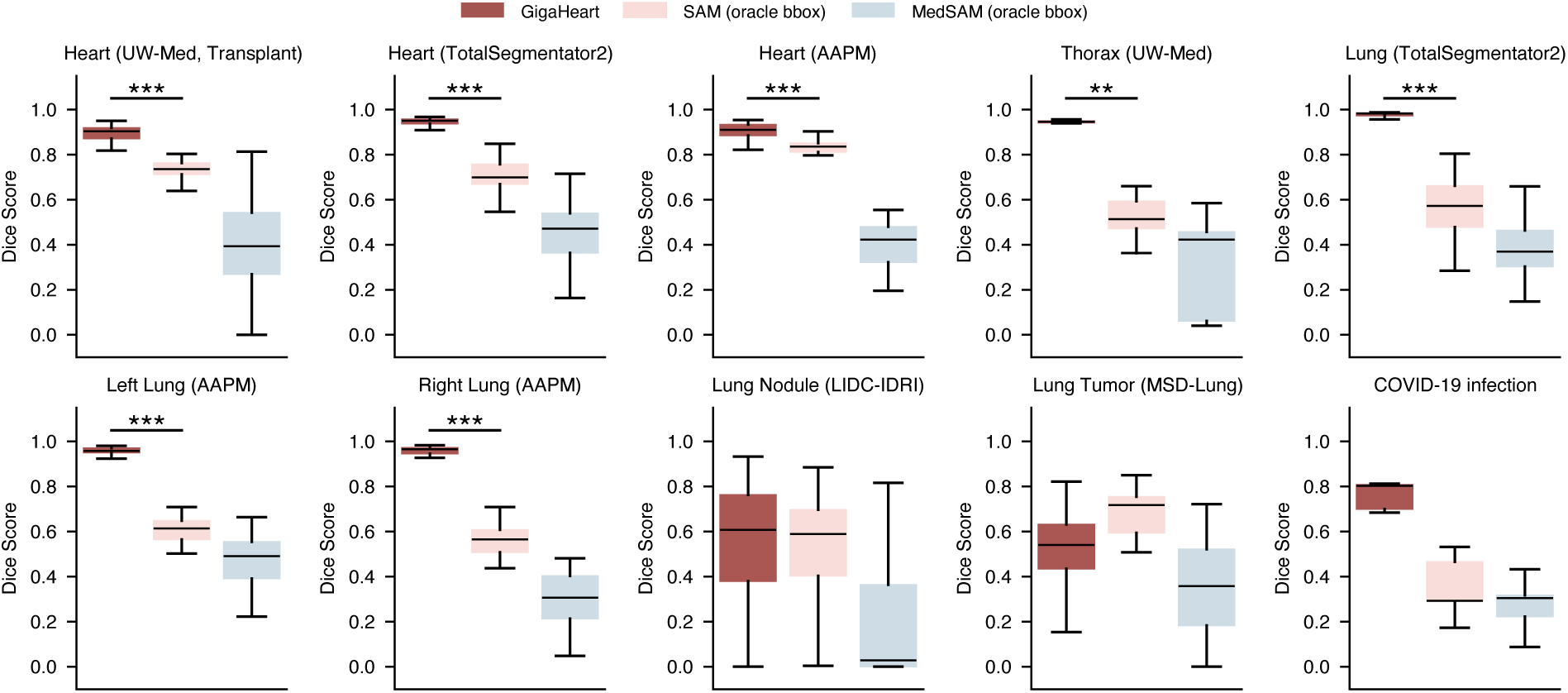
Box plot comparing *GigaHeart* with competing methods on cardiothoracic anatomy and abnormalities. The SAM and Med-SAM models use oracle bounding box as spatial prompt. Oracle bounding box is obtained from ground truth segmentation masks. ∗ indicates the significance level at which *GigaHeart* outperforms the best-competing method, with Wilcoxon test *p*-value< 1 × 10*^−^*^2^ for **, *p*-value< 1 × 10*^−^*^3^ for ***.

**Supplementary Figure 12:**
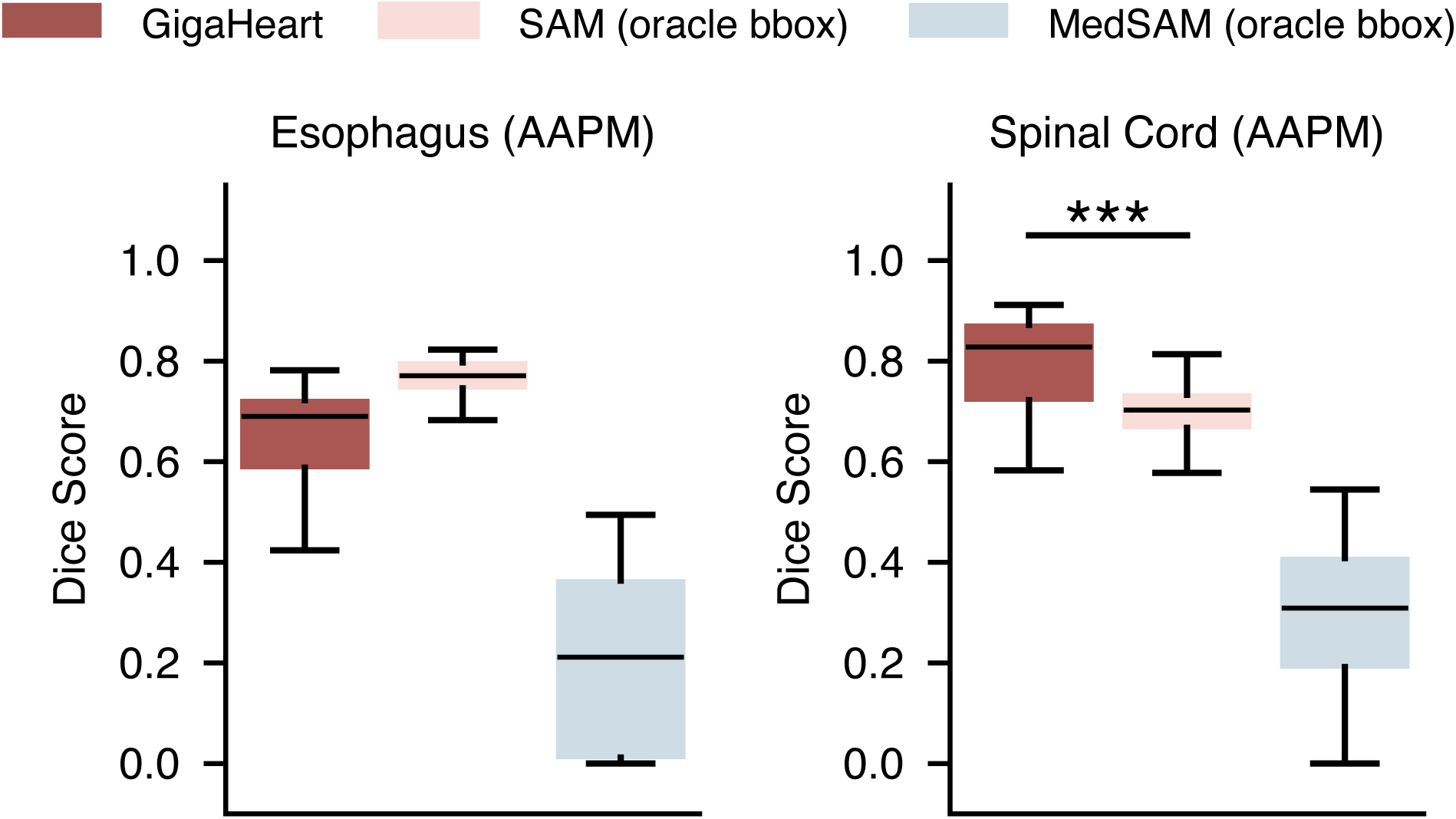
Box plot comparing *GigaHeart* with competing methods on segmenting the esophagus and spinal cord. The SAM and Med-SAM models use oracle bounding box as spatial prompt. Oracle bounding box is obtained from ground truth segmentation masks. ∗ indicates the significance level at which *GigaHeart* outperforms the best-competing method, with Wilcoxon test *p*-value< 1 × 10*^−^*^3^ for ***.

